# Erytra Blood Group Analyser and Kode Technology testing of SARS-CoV-2 antibodies among convalescent patients and vaccinated individuals

**DOI:** 10.1101/2021.08.26.21262219

**Authors:** Christof Weinstock, Willy A Flegel, Kshitij Srivastava, Sabine Kaiser, Hubert Schrezenmeier, Chrysanthi Tsamadou, Carolin Ludwig, Bernd Jahrsdörfer, Nicolai V Bovin, Stephen M Henry

**Author notes:** **Correspondence** Christof Weinstock, Institute of Clinical Transfusion Medicine and Immunogenetics, Helmholtzstrasse 10, 89081 Ulm, Germany. **Authorship contributions** CW and SK designed the study and acquired the data, CW analysed the data and wrote the manuscript draft. WAF and KS contributed to assay development and study design. NVB and SMH designed the C19-kodecyte assay and contributed to the manuscript. HS, CT, and BJ provided data from the CAPSID study, organised the VACCID study and wrote the manuscript. **Data availability statement** All data generated during and/or analyzed during the current study are available upon request by contact the corresponding author. **Disclaimer** The views expressed do not necessarily represent the view of the National Institutes of Health, the US Food and Drug Administration, the Department of Health and Human Services, or the U.S. Federal Government.

## Abstract

Surveillance of the severe acute respiratory syndrome coronavirus 2 (SARS-CoV-2) pandemic requires tests to monitor antibody formation and prevalence. We detected SARS-CoV-2 antibodies using red cells coated by Kode technology with short peptides derived from the SARS-CoV-2 spike protein. Such modified red cells, called C19-kodecytes, can be used as reagent cells in any manual or automated column agglutination assay. We investigated the presence of SARS-CoV-2 antibodies in 130 samples from COVID-19 convalescent plasma donors using standard manual technique, two FDA authorized ELISA assays and a virus neutralisation assay. The sensitivity of the C19-kodecyte assay was 88%, comparable to the anti-SP and anti-NCP ELISAs (86% and 83%) and the virus neutralisation assay (88%). The specificity of the C19-kodecyte assay was 90% (anti-SP 100% and anti-NCP 97%). Likewise, 231 samples from 73 vaccinated individuals were tested with an automated analyser and we monitored the appearance and persistence of SARS-CoV-2 antibodies. The C19-kodecyte assay is a robust tool for SARS-CoV-2 antibody detection. Automated blood group analyser use enables large-scale SARS-CoV-2 antibody testing for vaccination monitoring in population surveys.

## Introduction

In the COVID-19 pandemic, tests for virus RNA or virus particles enable the detection and isolation of infected individuals. The proportion of the population carrying antibodies following either infection or vaccination determines the herd-immunity. How long protective antibodies persist after infection or vaccination remains to be determined. Large-scale population screens will provide this valuable information and facilitate the surveillance during the pandemic.

Many platforms for SARS-CoV-2 antibody testing have been launched,^1^ typically requiring specialized liquid handling and reader devices for result evaluation. We recently developed C19-kodecyte reagent red cells suitable for routine manual and automated assays with the antiglobulin techniques available in most blood bank and hospital laboratories.^2,3^ C19-kodecyte reagent red cells can be prepared in any laboratory within 2 hours by inserting Kode Technology constructs into the membranes of blood group O red cells. The C19-kodecytes are thus coated with 15 amino acid-long peptides derived from the SARS-CoV-2 spike protein attached to the red cell membrane by a spacer and a lipid. The resultant reagent red cells are then tested against undiluted serum or plasma samples in any indirect antiglobulin platform.

As many immunohematology laboratories worldwide have automated blood group analysers, they are capable of large-scale testing and uniquely positioned to continuously survey their presumably healthy blood donor populations for COVID-19 immunity. Here we evaluated the C19-kodecyte assay in 130 convalescent plasma donors. The results were compared to established ELISA and a plaque reduction neutralisation assay.^1^ In addition, we transferred the C19-kodecyte assay onto an automated blood group analyser and evaluated 231 samples from a vaccination monitoring study.

## Materials and methods

### COVID-19-convalescent donor and control samples

Serum samples were sourced from blood donors who had recovered from mild to moderate PCR-confirmed COVID-19 disease and assessed as donors for convalescent plasma for a randomized prospective trial for treatment of patients with severe COVID-19 (CAPSID; EudraCT no. 2020-001310-38; ClinicalTrials.gov Identifier NCT04433910). All 130 samples were tested with the Euroimmun ELISA for antibodies directed against the spike protein (SP) and for antibodies against the nucleocapsid protein (NCP). In addition, 88 of these samples had been tested with the SARS-CoV-2 plaque reduction neutralisation test (PRNT)^1,4^ which detects the reduction of wild-type virus-induced cell culture plaques. The results of the PRNT are given as the titer of sample at which a reduction of the plaques by 50% (PRNT50) or 90 % (PRNT90) is observed. For the present study we used the PRNT50 results.

For negative controls, 38 serum samples were obtained from healthcare workers and their dependents (not known to have had COVID-19 or been vaccinated). Eleven of these control samples were included in a recently published study.^1^

### Plasma samples from SARS-CoV-2 vaccination screening programme

Informed consent was obtained and individuals were tested for antibodies against SARS-CoV-2 prior to and after vaccination. This study was approved by the ethics board of the University of Ulm (no. 488/20).

### C19-kodecytes

C19-kodecyte reagent red cells were prepared as previously described.^2^ In brief, the Kode constructs FSL-1147 and FSL-1255 were both dispersed in red cell stabilizer solution (ID-Cellstab 005650; Bio-Rad, München, Germany) at concentrations of 1.5 µmol/L and 2.5 µmol/L, respectively. The FSL-1147+1255 construct blend was incubated with washed packed group O red cells for 2 hours at 37 °C, then adjusted to 1 % using red cell stabilizer solution.

### C19-kodecyte assay

Serum samples from COVID-19 convalescent donors and controls were manually tested using Grifols DG antiglobulin and saline cards (no. 210342 and 210343, respectively; Grifols S.A., Barcelona, Spain). The cards were used according to the recommendations of the manufacturer. In brief, 25 µl of serum was incubated with 50 µl of 1 % C19-kodecytes in antiglobulin cards. All reactive samples were also tested against untreated cells (the same cells as used to make the kodecytes) in order to exclude reactivity caused by antibodies to natural red cell antigens. In addition, all samples were tested with C19-kodecytes in saline cards in order to determine the contribution of IgM to the reaction. Cards were incubated at 37 °C for 30 min, centrifuged in a DG Spin centrifuge (Grifols S.A., Barcelona, Spain), and the reactions were graded according to the scheme shown in Fig 1.

**Fig. 1:**
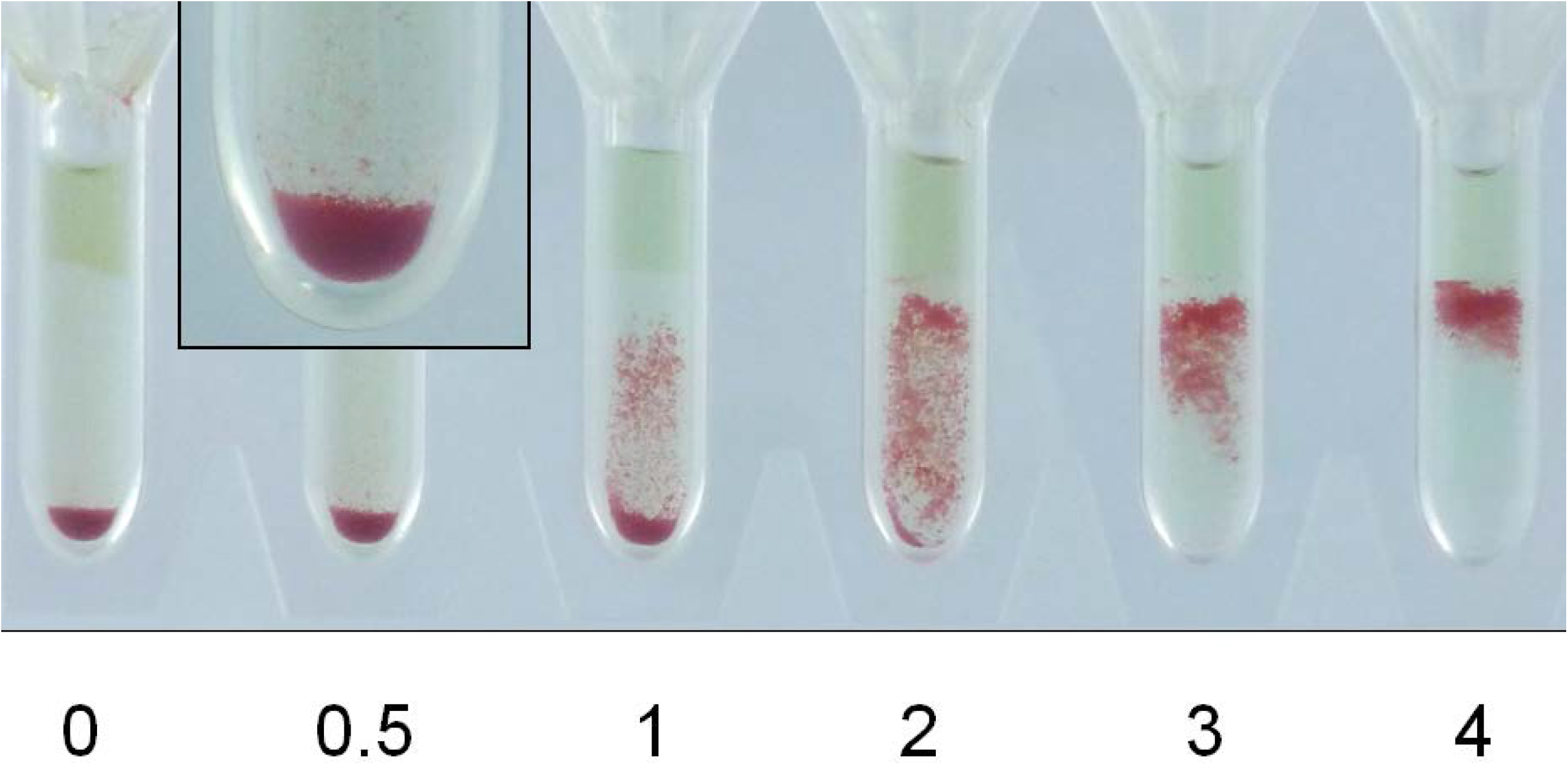
Grading of the reaction strength used for the C19-kodecyte assay. C19-kodecyte agglutination grades are shown for gel cards with antiglobulin. Grade 0: no cells above the pellet; grade 0.5: only very few cells above the pellet (a magnification of the reaction is inserted); grade 1: cells are disseminated in the column, most of them located in the lower third; grade 2: cells are disseminated in the column; grade 3: most cells are located in the upper third of the column, no cells at the bottom; grade 4: cells form a band at the top of the column.

### Automation of the C19-kodecyte assay

Blood samples from the vaccinated individuals were tested with the Grifols Erytra Automated System (Grifols S.A., Barcelona, Spain). An antibody screening test with antiglobulin cards was done against C19-kodecytes, untreated control cells (being the same cell as used to make the kodecytes), a 3-cell pool of antibody screening reagent cells and an autocontrol (patient’s own cells). After completion of the tests, the gel cards were visually reviewed and the reactions were graded according the scheme shown in Fig 1. In all cases, the visual grading was consistent with the grading of the Erytra.

### Statistics

The sensitivity of the assays was calculated as the proportion of convalescent samples which gave a positive test result. Specificity was calculated as the proportion of control samples which gave a negative test result.

The positive predictive value of an assay was calculated by dividing the number of positive convalescent samples by the sum of positive convalescent samples and positive samples from controls. The negative predictive value of an assay was calculated by dividing the number of negative samples from controls by the sum of negative samples from controls and negative convalescent samples.

## Results

### Performance of the C19-kodecyte assay

Serum samples from 130 convalescent plasma donors and 38 controls were tested with the C19-kodecyte assay. In the convalescent donor samples, 114 reacted positive with the C19-kodecytes in antiglobulin cards and 9 of these samples also reacted positive in saline cards, indicating the presence of IgM (Fig 2). Four of the 38 control samples reacted positive with C19-kodecytes in antiglobulin cards (C19-kodecyte reaction grades of 1, 1, 2, 2), with none being reactive in saline cards.

**Fig. 2:**
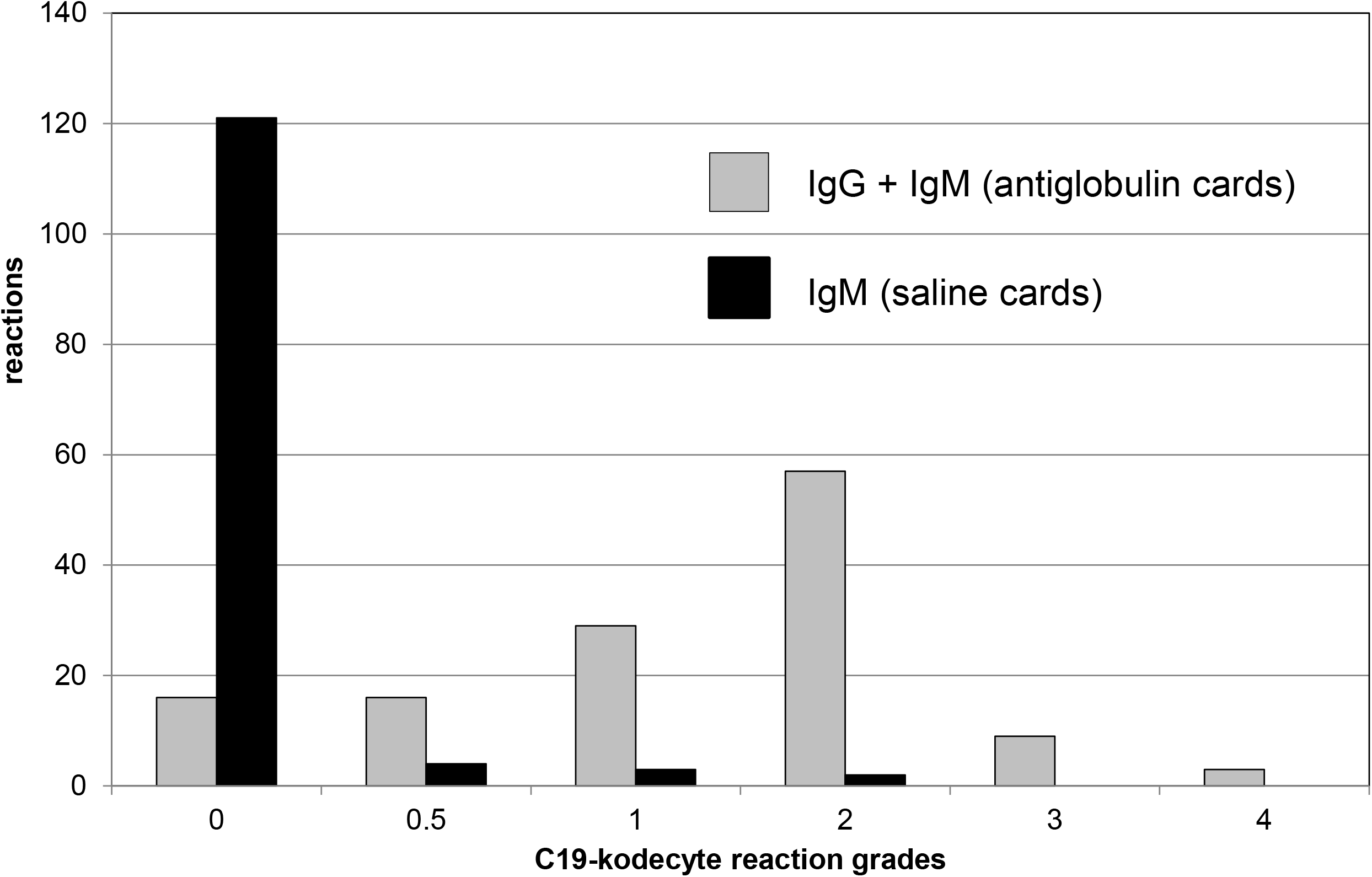
Results for 130 serum samples from convalescent plasma donors. The sera were tested by the C19-kodecyte assay using antiglobulin (grey bars) or saline (black bars) gel cards.

All samples were tested with the Euroimmune ELISA. The convalesecent donor samples reactive with the anti-SP-ELISA were additionally tested with the virus neutralization assay. The results were compared to the results of the C19-kodecyte assay. The sensitivity of the C19-kodecyte assay was 88 % (Table I), compared to 86 % for the anti-SP IgG ELISA, 83 % for the anti-NCP IgG ELISA, and 88 % for the virus neutralization assay. The specificity of the C19-kodecyte assay was 90 %, while specificity of the ELISA for detecting anti-SP IgG and anti-NCP IgG was 100 % and 97 %, respectively.

**Table I.** Comparison of the C19-kodecyte assay with ELISA and virus neutralisation assays.

|  | C19-kodecyte | ELISA |  | virus<br>neutralization |
| --- | --- | --- | --- | --- |
|  |  | spike<br>protein<br>IgG | nucleocapsid<br>protein, IgG |  |
| Convalescent donors (n) | 130 | 130 | 130 | 88 |
| Positive results | 114 | 112 | 108 | 77 |
| Negative results | 16 | 18 | 22 | 11 |
| Negative controls (n) | 38 | 38 | 38 | NT |
| Negative results | 34 | 38 | 37 |  |
| Positive results | 4 | 0 | 1 |  |
| Assay performance |  |  |  |  |
| Sensitivity (%) | 88 | 86 | 83 | 88 |
| Specificity (%) | 90 | 100 | 97 | NA |
| Positive predictive value (%) | 97 | 100 | 99 | NA |
| Negative predictive value (%) | 68 | 68 | 63 | NA |

ELISA, enzyme-linked immunosorbent assay (Euroimmun); NT, not tested; NA, not applicable.

### Comparison of the C19-kodecyte assay with the ELISA and the PRNT

C19-kodecyte assay reaction grades (Fig 1), which semi-correlate with antibody levels,^5^ were compared with ELISA optical densities (OD). The C19-kodecyte assay grades of the 130 convalescent donors samples correlated well with the means of the ELISA ratios for anti-SP IgG (R^2^ = 0.95, Fig 3, Panel A) and for anti-NCP IgG (R^2^ = 0.96, Fig 3, Panel B). Of the 88/130 samples tested with the virus neutralisation assay, the number of positive C19-kodecyte results also correlated with the virus neutralization assay results (Fig 3, Panel C). These same 88 serum samples were analysed with the anti-SP IgG ELISA and correlated well to the virus neutralization assay results (R^2^ = 0.88). Of note, only samples reactive with the ELISA had been selected for testing against the virus neutralization assay, which explains the lack of negative ELISA results (Fig. 3, Panel D).

**Fig. 3:**
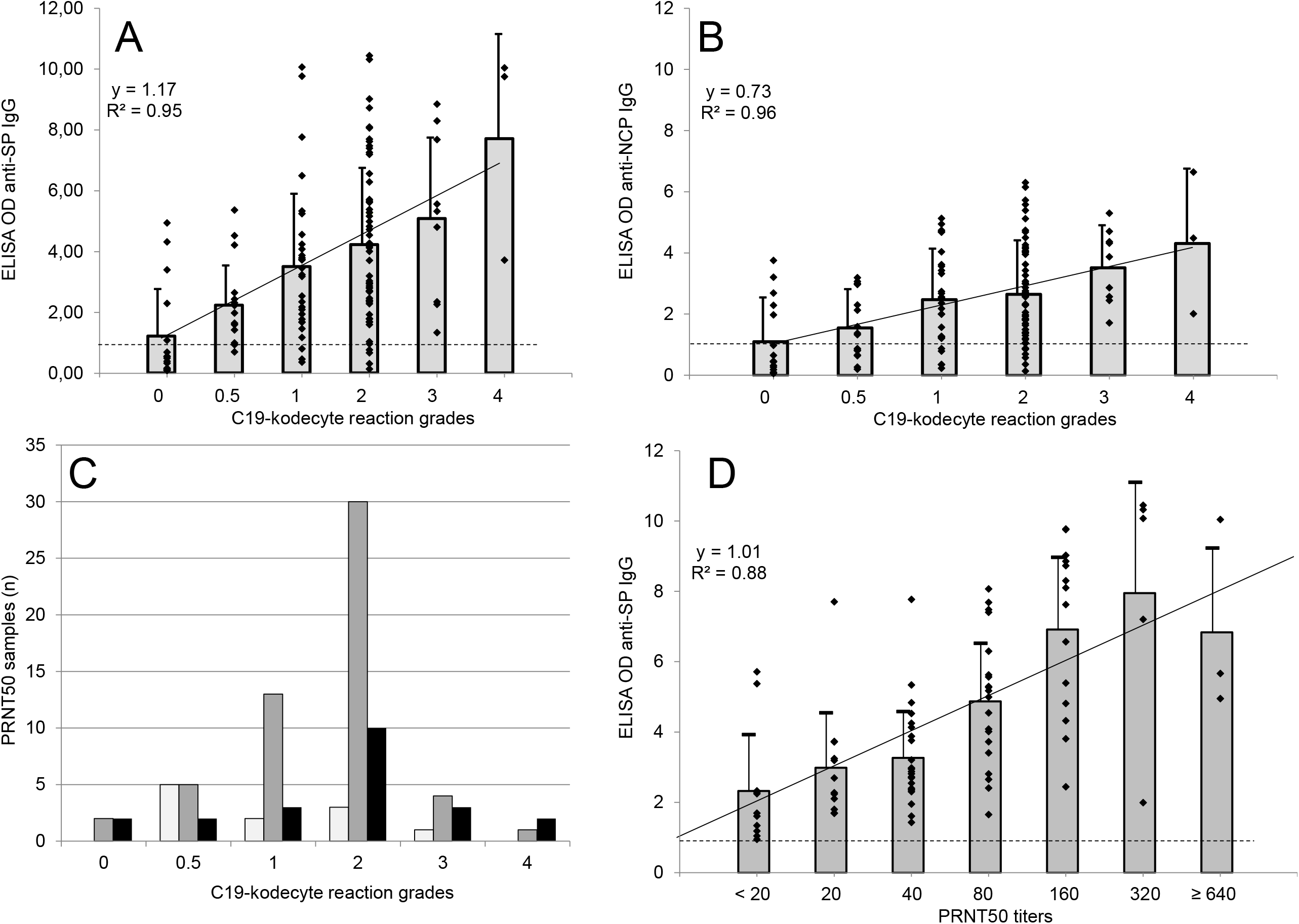
Comparison of the C19-kodecyte assay with the ELISA and the PRNT. Serum samples from 130 SARS-CoV-2 convalescent donors. Samples were grouped according to their grades in the C19-kodecyte assay and compared to the results of the ELISA for anti-SP IgG (A) and anti-NCP IgG (B). The reaction strength grading of the C19-kodecyte assay are given on the x-axis, the optical density (OD) ratio results of the ELISA are given on the y-axis, each diamond represents one serum sample. The bars indicate the mean OD ratio, and the T-bars indicate the standard deviation. The dotted line represents the cut-off (0.8 OD ratio) (C) 88 of the 130 serum samples were tested against the virus neutralization assay and compared with the results of the C19-kodecyte assay. Samples were grouped according to the PRNT50 titer results: < 20 (light grey bars), between 20 and 80 (grey bars), and ≥ 160 (black bars). (D) For comparison, the results of the anti-SP IgG ELISA of the 88 samples which also were tested with the virus neutralization assay were analysed. The results were grouped according to the titres given by the PRNT50.

A total of 76% (99/130) convalescent donor samples reacted positive with all three methodologies, 6% (8/130) were negative with all three methodologies, while 18% (23/130) were discordant, with at least one assay being negative. Of these 23 discordant results (Table II) 15 were positive with C19-kodecytes, of which 11 were either positive with the anti-SP (n=7) or anti-NCP (n=12) ELISA. Eight samples were negative with C19-kodecytes and positive by one or more to the ELISA assays. Five samples were negative with C19-kodecytes (which is an anti-SP assay) and positive with the anti-SP ELISA while in contrast 7 samples were positive with C19-kodecytes and negative with the anti-SP ELISA

**Table II.** Clustering of discordant negative results between methodologies for convalescent samples.

| Convalescent | Methodologies |  |  |
| --- | --- | --- | --- |
| Samples (n) | C19-kodecytes | ELISA anti-SP | ELISA anti-NCP |
| 8 | + | + | - |
| 3 | + | - | + |
| 4 | + | - | - |
| 3 | - | + | + |
| 3 | - | - | + |
| 2 | - | + | - |

ELISA, enzyme-linked immunosorbent assay; SP, spike protein; NCP, nucleocapsid protein.

### Automation of the C19-kodecyte assay

A total of 231 blood samples from 73 vaccinated individuals were tested with the Erytra Automated System. The routine antibody screening programme was employed, which encompasses 3 reagent cells and an autocontrol. The automated grading was in accordance with the grading as defined in Fig 1 and none of the results were manually edited. Data on the participants are shown in Table III. The results of 26 study participants who had an initial negative first sample result and then became positive are shown in Fig 4. All 26 individuals were antibody positive by day 96 post vaccination, with the majority (18/26, 69%) being antibody positive by day 44. The majority of reactions grades for immunised individuals (19/26, 73 %) was grade 2+ or greater, with only 7 individuals having 1+ or weaker grades. The 8 samples which had an initial negative result but had not yet become assay positive ranged from 22 to 101 days post vaccination.

**Table III.**
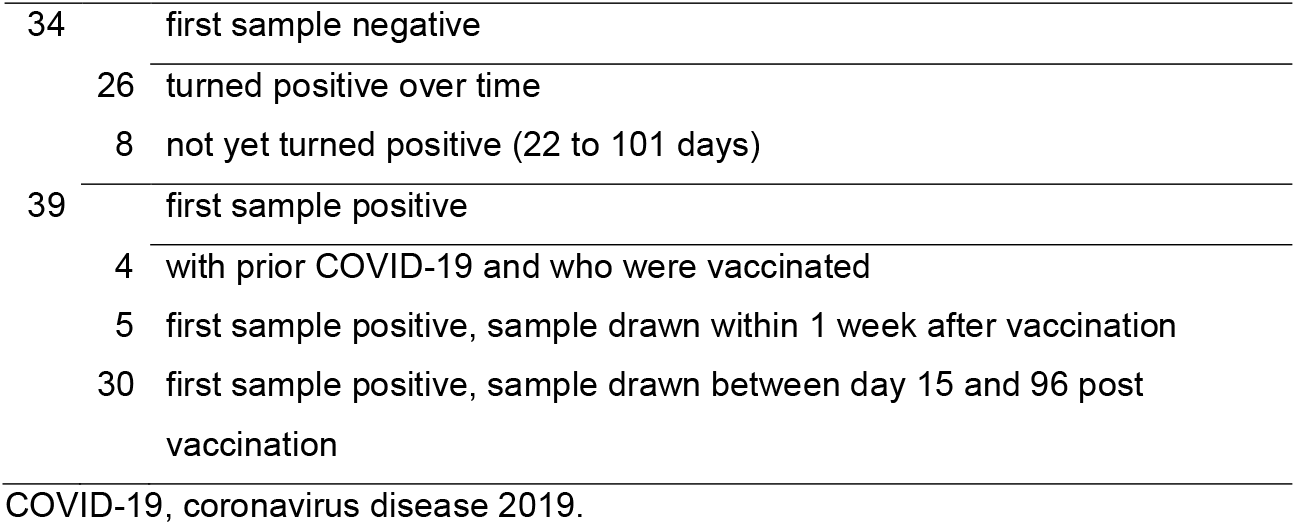
Interim data of 73 participants of an ongoing vaccination study.

**Fig. 4:**
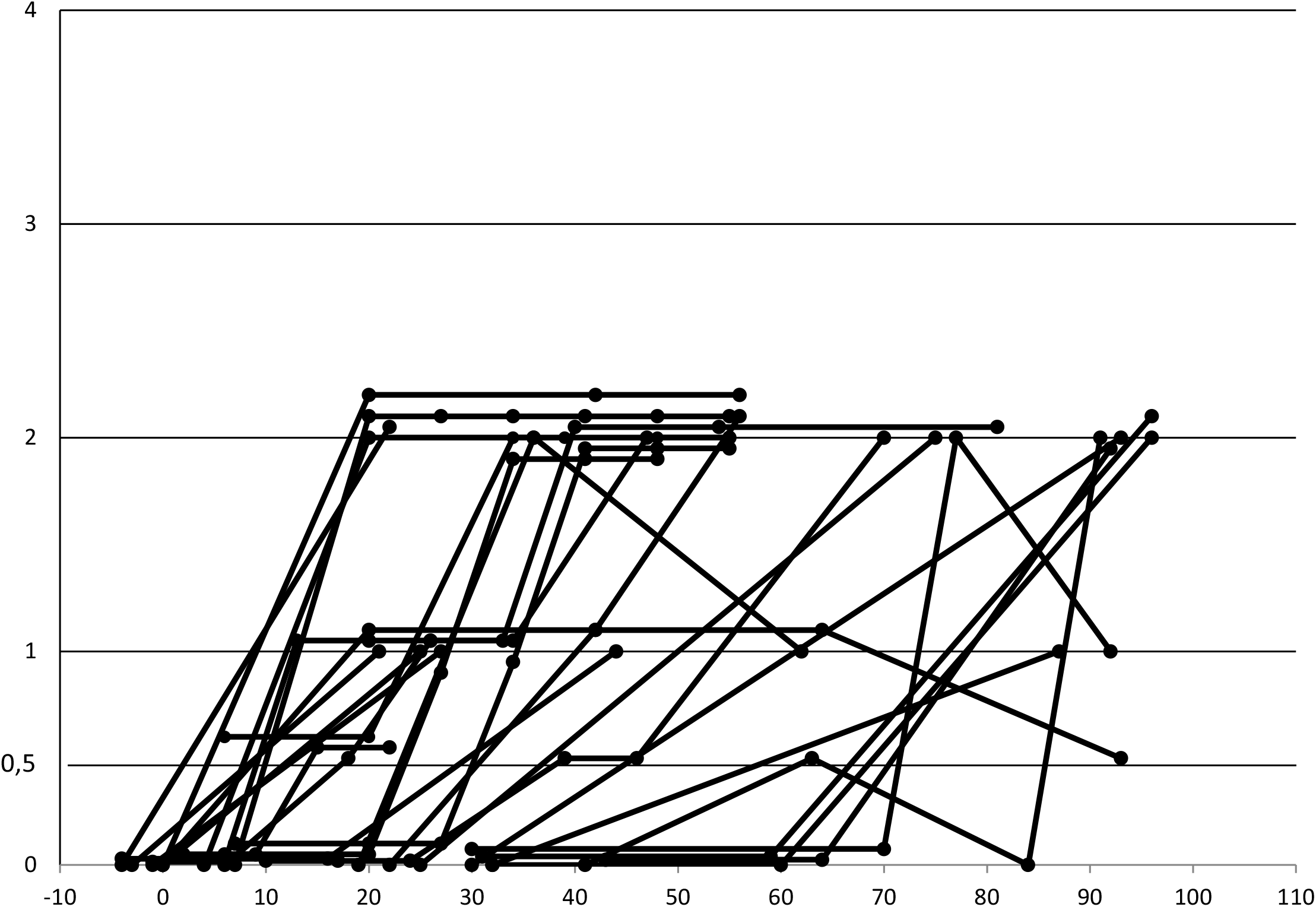
C19-kodecyte grades over time of 26 vaccinated individuals without known prior exposure to SARS-CoV-2. These individuals had a negative first sample and developed antibodies over time.

## Discussion

In this study, we further investigated the novel C19-kodecyte assay designed for the detection of SARS-CoV-2 antibodies. Red cells were coated with short peptides derived from the SARS-CoV-2 spike protein.^2,6^ These red cells laden with artificial SARS-CoV-2 spike antigens (C19-kodecytes) behave like normal red cells bearing blood group antigens in routine antibody screening assays. We first investigated the performance of the C19-kodecyte assay against convalescent plasma donor samples. We then transferred the C19-kodecyte assay onto the Erytra Automated System, because automated processing would facilitate testing of large numbers of blood samples required for vaccination monitoring during the SARS-CoV-2 pandemic.

The preparation of the C19-kodecytes is simple and only involves the incubation of washed packed red cells with specifically designed peptides (Kode constructs) for 2 hours. The constructs spontaneously self-assemble into the cell membrane. No special equipment or training of the laboratory staff is required for implementation of the C19-kodecyte assay, because the reactions observed are typical agglutination reactions, routinely observed in red cell serology testing.

Samples from convalescent donors were tested against a virus neutralisation assay and against two ELISA platforms for the detection of SARS-CoV-2 antibodies. We compared the C19-kodecyte assay (a spike protein anti-IgG assay with limited reactivity to IgM) with the results from these other platforms. Sensitivity (the estimated proportion of subjects with the target condition in whom the test is positive), positive predictive value, and negative predictive value were comparable for the four assays. The calculated specificity (the estimated proportion of subjects without the target condition in whom the test is negative) of the C19-kodecyte assay was lower than that of the ELISA platforms with 4 of 38 samples from individuals without known SARS-CoV-2 infection reacting positive. Although these reactions were caused by IgG binding to the C19-kodecyte reagent red cells, it could not be clarified whether the reaction of these four samples were nonspecific (an unknown specificity reacting against the peptide), or whether the donors had an asymptomatic SARS-CoV-2 infection, or had cross reacting antibodies from previous infections with seasonal alpha and beta-corona viruses.^7-9^ However, this rate was similar to the 91% specificity rate reported previously for the Grifols platform.^2,6^ The samples from negative controls had not been tested with the virus neutralization assay, so a comparison was not possible.

Although the majority of convalescent samples reacted positive with all three assays, there were also 31 samples that unexpectedly reacted negative with one or more assays. Eight samples were negative with all three assays suggesting that the level of antibodies in these samples were below the detection threshold. C19-kodecytes reacted positive with 15 of these 23 discordant samples (compared with 13/23 for anti-SP ELISA and 9/23 for anti-NCP ELISA) and therefore was unable to detect antibody in only 8 samples that were positive by either or both of the ELISA assays.

This divergent reactivity of the C19-kodecyte assay with the ELISA assays has two major probable causes. First, the polyclonal antibody response after infection can differ between individuals, i.e. one individual does not produce antibodies against the same epitopes of the virus as another individual. Therefore, individuals lacking or having lower levels of the antibodies specific for the target used in a specific assay may react negative, while reacting positive with another assay bearing a different target. Secondly, it should be noted that it is the cumulative result of all bound antibodies which is measured in an assay. The C19-kodecyte assay only utilises two linear mono epitopes (MEps) from a domain of the spike protein which is located closely to the virus membrane,^10^ whereas the ELISA assays utilise the S1 domain of the spike protein (SP), or a modified nucleocapsid protein (NCP), respectively. ELISA assays with large recombinant proteins are therefore representative of many linear and conformational epitopes and can therefore cumulatively capture many different antibodies (including non-specific antibodies). In contrast C19-kodecytes, represented by only 2 linear MEps, can capture a very limited range of antibodies. These assay differences have both positive and negative consequences, and must be balanced (primarily through selection of MEps and their relative concentration(s) on the outside of the cell). The presentation of a limited number of precisely selected epitopes, and exclusion of undesired epitopes (which are unavoidable on recombinant proteins), allows for the use of undiluted serum in kodecyte assays with a consequent potential increase in sensitivity. However, restriction of epitope selection on kodecytes also means a reduction in sensitivity due to the loss of a cumulative effect of detecting multiple different antibodies as occurs with recombinant proteins in ELISA assays.

The assay present in this paper is a beta version and ultimately as new knowledge is obtained, the C19-kodecyte assay can be adapted and sensitivity and specificity improved by exchanging or adding new MEps (and adjusting their relative concentrations). This also gives the opportunity for any kodecyte assay to be tuned to bearing the most clinically relevant MEps, an opportunity not readily available to most recombinant protein methodologies.

For surveillance and control of the SARS-CoV-2 pandemic, it is necessary to know the current immunity status of the population. Early data on SARS-CoV-2 immunity suggested a rapid decline of the antibody levels,^11,12^ raising questions on the risk of re-infection of convalescents, while others found longer lasting antibody persistence.^13^ Testing of large numbers of convalescents and vaccinated people for the collection of large data is required in order to draw meaningful conclusions. The C19-kodecyte assay can support such efforts because it is based on the mass-screening indirect antiglobulin technique which is established in all medical laboratories performing immunohematological investigations. We evaluated the C19-kodecyte assay on the Grifols Erytra blood group analyser and tested 231 samples from 73 participants of a vaccination study in parallel with patient blood grouping and cross matching, without interference with the laboratory routine. The C19-kodecyte results showed appearance and persistence of antibodies against SARS-CoV-2 in vaccinated individuals. Kodecyte have been previously established to be semi-quantitative,^5^ and it can be seen that the C19-kodecyte assay also appears to be semi-quantitative, in that serological C19-kodecyte grades are indicative of absent, low and high levels of antibody. Post vaccination the C19-kodecyte grade of most individuals rose to 1+ or 2+. Although not yet established, C19-grades of 2+ maybe indicative of fully immunised status while those with grades of 1+ or less may not yet be optimally immunised. This could be important for future trials in determining the immunisation status/risk of a population.

The C19-kodecyte assay does not require any special equipment, it can be performed manually, although the use of an automated blood group analyser increases the number of samples which can be tested. The Erytra Automated System uses column agglutination cards with 8 reaction columns and is capable of routinely processing at least 50 cards per hour. If just screening for SARS-CoV-2-antibody test was to be done with an autocontrol (i.e 4 samples per card), then 200 samples could be tested per hour, especially in times when the analyser otherwise is idle. However, as a positive autocontrol result (being unmodified cells used to make kodecytes) is due to the presence of natural red cell antibodies, the autocontrol need not be done when testing blood donor populations who are already screened for red cell antibodies. Therefore, when screening blood donors 8 samples could be tested per column card, allowing for potentially 400 tests per hour.

As the reagent costs for the preparation of the kodecytes are below 0.10 € per test, and implementation is easy, this assay is potentially a valuable tool for the efforts of monitoring population immunity status in the SARS-CoV-2 pandemic.

## Data Availability

All data generated during and/or analyzed during the current study are
available upon request by contact the corresponding author.

## Acknowledgements

Part of this work has been published in abstract form at the online meeting of the German Society for Transfusion Medicine and Immunohaematology (DGTI), 22 September 2021. WAF was supported by the Intramural Research Program (project ID ZIC CL002128) of the NIH Clinical Center at the National Institutes of Health. Development and supply of FSL constructs was supported the New Zealand Ministry of Business, Innovation & Employment COVID-19 Innovation Acceleration Fund, contract CIAF 0490.

